# GWAS reveals Genetic Susceptibility to Air Pollution-Related Asthma Exacerbations in Children of African Ancestry

**DOI:** 10.1101/2024.05.29.24307906

**Authors:** Jelte Kelchtermans, Michael E. March, Frank Mentch, Yichuan Liu, Kenny Nguyen, Hakon Hakonarson

## Abstract

**Background:** The relationship between ambient air pollution (AAP) exposure and asthma exacerbations is well-established. However, mitigation efforts have yielded mixed results, potentially due to genetic variability in the response to AAP. We hypothesize that common single nucleotide polymorphisms (SNPs) are linked to AAP sensitivity and test this through a Genome Wide Association Study (GWAS).

**Methods:** We selected a cohort of pediatric asthma patients frequently exposed to AAP. Patients experiencing exacerbations immediately following AAP spikes were deemed sensitive. A GWAS compared sensitive versus non-sensitive patients. Findings were validated using data from the All of Us program.

**Results:** Our study included 6,023 pediatric asthma patients. Due to the association between AAP exposure and race, GWAS analysis was feasible only in the African ancestry cohort. Seven risk loci reached genome-wide significance, including four non-intergenic variants. Two variants were validated: rs111970601 associated with sensitivity to CO (odds ratio [OR], 6.58; PL=L1.63L×L10−8; 95% CI, 3.42-12.66) and rs9836522 to PM2.5 sensitivity (OR 0.75; PL=L3,87 ×L10−9; 95% CI, 0.62-0.91).

**Interpretation:** While genetic variants have been previously linked to asthma incidence and AAP exposure, this study is the first to link specific SNPs with AAP-related asthma exacerbations. The identified variants implicate genes with a known role in asthma and established links to AAP. Future research should explore how clinical interventions interact with genetic risk to mitigate the effects of AAP, particularly to enhance health equity for vulnerable populations.

**What is already known on this topic:** The relationship between ambient air pollution (AAP) exposure and asthma exacerbations is well-established. However, efforts to mitigate the impact of AAP on children with asthma have yielded mixed results, potentially due to genetic variability in response to AAP.

**What this study adds:** Using publicly available AAP data, we identify which children with asthma experience exacerbations immediately following spikes in AAP. We then conduct a Genome Wide Association Study (GWAS) comparing these patients with those who have no temporal association between AAP spikes and asthma exacerbations, identifying several Single Nucleotide Polymorphisms (SNPs) significantly associated with AAP sensitivity.

**How this study might affect research, practice, or policy:** While genetic variants have previously been linked to asthma incidence and AAP exposure, this study is the first to link specific SNPs with AAP-related asthma exacerbations. This creates a framework for identifying children especially at risk when exposed to AAP. These children should be targeted with policy interventions to reduce exposure and may require specific treatments to mitigate the effects of ongoing AAP exposure in the interim.

## Introduction

While inequity in asthma incidence and mortality in the United States is well documented, addressing the factors that drive this inequity has proven challenging.(1) One of these factors is Ambient air pollution (AAP) exposure. AAP exposure is associated with asthma incidence, has been shown to trigger asthma exacerbations, and is closely linked to racial and ethnic status in the United States. (2–5)

The clinical impact of AAP is determined by two main factors: the level and duration of pollution exposure itself (‘AAP exposure’), and the individual susceptibility to this exposure (‘AAP sensitivity’). Seminal work in the field has detailed how AAP exposure is linked to clinical outcomes such as asthma incidence and severity. (3, 6) However, considerably less is known about how individual AAP sensitivity impacts clinical outcomes, in particular exacerbations. This knowledge gap has likely contributed to inconsistent results in prior studies evaluating strategies to prevent asthma exacerbations in pediatric patients exposed to AAP. (7, 8)

If we can identify genetic drivers of AAP sensitivity, we may be able to identify at risk patients and high-value therapeutic targets. While the long-term imperative is clearly to reduce AAP exposure and remove inequities in AAP exposure, strategies to mitigate the impact of this exposure are immediately needed. In the presented work, we sought to achieve this by integrating clinical, genomic, and environmental data to identify patients sensitive to spikes in AAP and then complete a genome wide association study (GWAS) to identify single nucleotide polymorphisms (SNPs) associated with this sensitivity. Of note, this study drew pediatric patients with asthma frequently exposed to ambient air pollution from a larger cohort of pediatric patients with asthma. We previously assessed air pollution sensitivity in the larger asthma cohort and published associated clinical characteristics. (9)

## Methods

### Population

Our population was drawn from patients with asthma enrolled in the Center of Applied Genomics biobank at the Children’s Hospital of Philadelphia. Cohort characteristics were described previously. (9) This study used only data from an existing deidentified database, did not involve interactions with human subjects, and posed minimal risk to privacy and confidentiality and as such is not considered human subject’s research.

### EPA data

We selected a sub-group of patients frequently exposed to poor air quality using annual Air Quality Index (AQI) data. (10) We calculated the ratio of number of days where air quality was not classified as ‘Good’ over the total number of days with air quality available over the past 15 years (2005–2020). Patients living in ZIP codes where this ratio was above the 75^th^ percentile for all ZIP codes in our sample were retained for further analysis.

Next, we used one of two methods to address short term variation in AAP components. For particulate matter 2.5 (PM2.5) and Ozone (O3) daily AQI data for days were these pollutants were used to calculate overall AQI were used. (10) Since CO, NO2, PM10 and SO2 are rarely the pollutants with the highest associated AQI, we modelled exposure to these pollutants for patients living in Philadelphia County. Patients were considered sensitive to a specific AAP component if they experienced at least one asthma exacerbation preceded by a spike at least two standard deviations above baseline exposure on the day of exacerbation or three preceding days. Additional detail on EPA data processing and baseline exposure determination is provided in the online data supplement.

### GWAS

Genotype data were generated on genotyping array families from Illumina. Ancestry was assigned based on the results of principal component analysis (PCA). After splitting of ancestries, ancestry specific PCAs were performed. Additional detail on the processing and quality control of genotyping data is provided in the online data supplement.

### Statistical analysis

SAIGE was used for association testing. (11) Imputed genotypes were converted to Plink format for analysis. (12) A null logistic mixed model was fitted for each ancestry using the variants from the ancestry specific PCAs, including as covariates the 10 principal components, subject age and sex, and a covariate to adjust for the imputation file set in which each array was imputed. Sensitivity to each AAP component was treated as a binary trait and analyzed with saddle point approximation. Analysis was limited to SNPs with a minor allele count of at least 20. A p-value < 5 × 10^−8^ was used as the cut-off for genome wide significance.

### In silico validation

A literature search was completed for variants that met a genome wide significance threshold and the GTEX database was queried using the Metasoft tool (See acknowledgement section). (13) We also assessed the potential impact of these variants with the Ensembl Variant Effect Predictor tool. (14) For non-intergenic variants, the most likely associate gene was assigned with the Open Targets Genetics variant to gene tool and HaploReg was queried to assess regulatory potential. (15–17)

### External validation

The All of Us cohort was used for external validation. (18) Asthma was defined in line with our own asthma phenotype with patients required to have at least two clinical encounters with an asthma related diagnosis and at least one asthma related prescription. (9) Asthma exacerbations were defined by diagnostic codes with a fourteen-day lockout period between exacerbations. The All of Us cohort uses survey data to determine home 3-digit ZIP code and time at current address. With the available data, the interval from 01/01/2013 to 12/31/2017 was found to maximize the number of subjects with stable address and as such, patients with asthma, no missing ZIP code or AAP data, and at least one exacerbation in this interval were included. Publicly available EPA data was used to determine mean AAP concentration in home ZIP code during day of exacerbation and three preceding days. A t-test was used to compare this mean between patients with and without the SNPs with genome wide significance in our primary analysis. If this t-test revealed a significant difference, a linear regression model correcting for race, ethnicity, sex at birth, and smoking status (ever smoker as opposed to never smoker), as well as clustering at the patient level was used to correct for significant confounders. For variants that only occur within a specific genetic population, only controls of the same genetic ancestry were included for comparison.

## Results

### Population

A total of 8,125 pediatric with asthma were identified (**e-Table 1**). The 75^th^ percentile for occurrence of AQI >50 was 41% of days, leading patients living in ZIP codes with more frequent occurrences of poor air quality to being classified as experiencing Frequent AAP-Exposure (**Fig. 1**). Overall, 6,023 patients lived in Frequent AAP-Exposure areas (**Table 1**). We obtained AQI based on Ozone for 21,817 exacerbations and based on PM2.5 for 25,480 exacerbations. We modelled exposure to CO for 10,925 exacerbations, to NO2 for 15,977 exacerbations, to PM10 for 8,716 exacerbations and to SO2 for 16,452 exacerbations. The mean AQI was 53 (with standard deviation [Sd] 27) based on Ozone and 54 (Sd 16) based on PM2.5. The mean modelled AAP concentration was 0.33 parts per million (Sd 0.21) for CO, 16.51 parts per billion (Sd 8.29) for NO2, 23.29 microgram per cubic meter (Sd 11.50) for PM10, and 1.97 parts per billion (Sd 2.04) for SO2.

**Figure 1.**
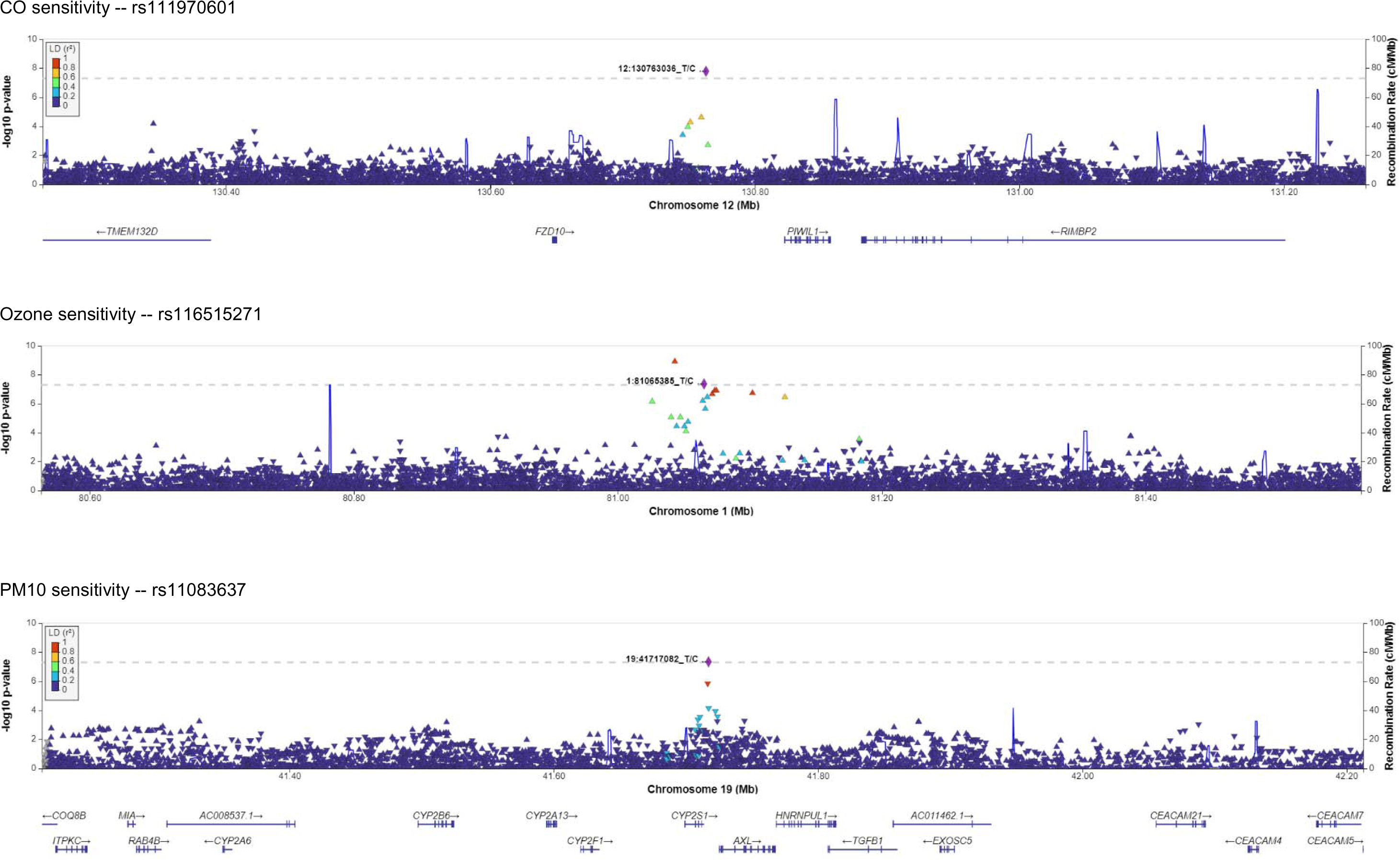
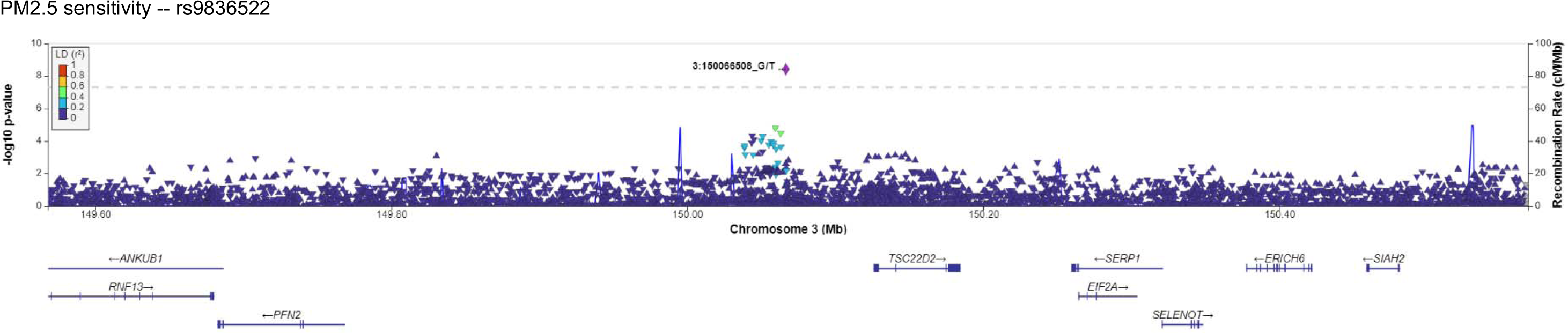
Study design: 8,125 pediatric patients with asthma were identified. Of this cohort, 6,023 patients lived in areas with frequent AAP-exposure and were included in the study.

**Table 1.** Population characteristics.

|  | Population |
| --- | --- |
| Number of patients | 6023 |
| Sex = Male (%) | 3406 (56.5) |
| Diagnosis (%) |  |
| Not Specified | 381 (6.3) |
| Mild Asthma | 3512 (58.3) |
| Moderate Asthma | 1822 (30.3) |
| Severe Asthma | 308 (5.1) |
| Documented allergy = Yes (%) | 4894 (81.3) |
| Number of Exacerbations (mean (SD)) | 4.25 (5.64) |
| Race – As documented in EMR (%) |  |
| Asian American | 86 (1.4) |
| Black or African American | 4596 (76.3) |
| White | 961 (16.0) |
| Other or missing | 380 (6.3) |
| Family History of atopic disease = Yes (%) | 4351 (72.2) |
| Obese = Yes (%) | 2030 (33.7) |
| History of RSV = Yes (%) | 603 (10.0) |
| History of Rhinovirus = Yes (%) | 620 (10.3) |
| Gestational Age (%) |  |
| Extremely preterm | 93 (1.5) |
| Very preterm | 153 (2.5) |
| Late preterm | 584 (9.7) |
| Early term | 805 (13.4) |
| Full term | 4388 (72.9) |
| Household income - USD (mean (SD)) | 40,246 (21,376) |

### Sensitive vs non-sensitive

Based on temporal association between asthma exacerbation and spike in AAP exposure, 1,282 patients were classified as sensitive to CO (39% of patients with CO data available), 1,471 to NO2 (36%), 671 to PM10 (27%), 1,905 to SO2 (45%), 1,504 to Ozone (27%), and 2,219 to PM2.5 (37%) (**e-Table 2**). For every pollutant, there was a significant difference in asthma severity between sensitive and non-sensitive patients (**Fig. 2**). Sensitive patients were consistently more likely to have a documented allergy, family history of atopic disease, and polymerase chain reaction confirmed rhinovirus or RSV infection.

**Figure 2.**
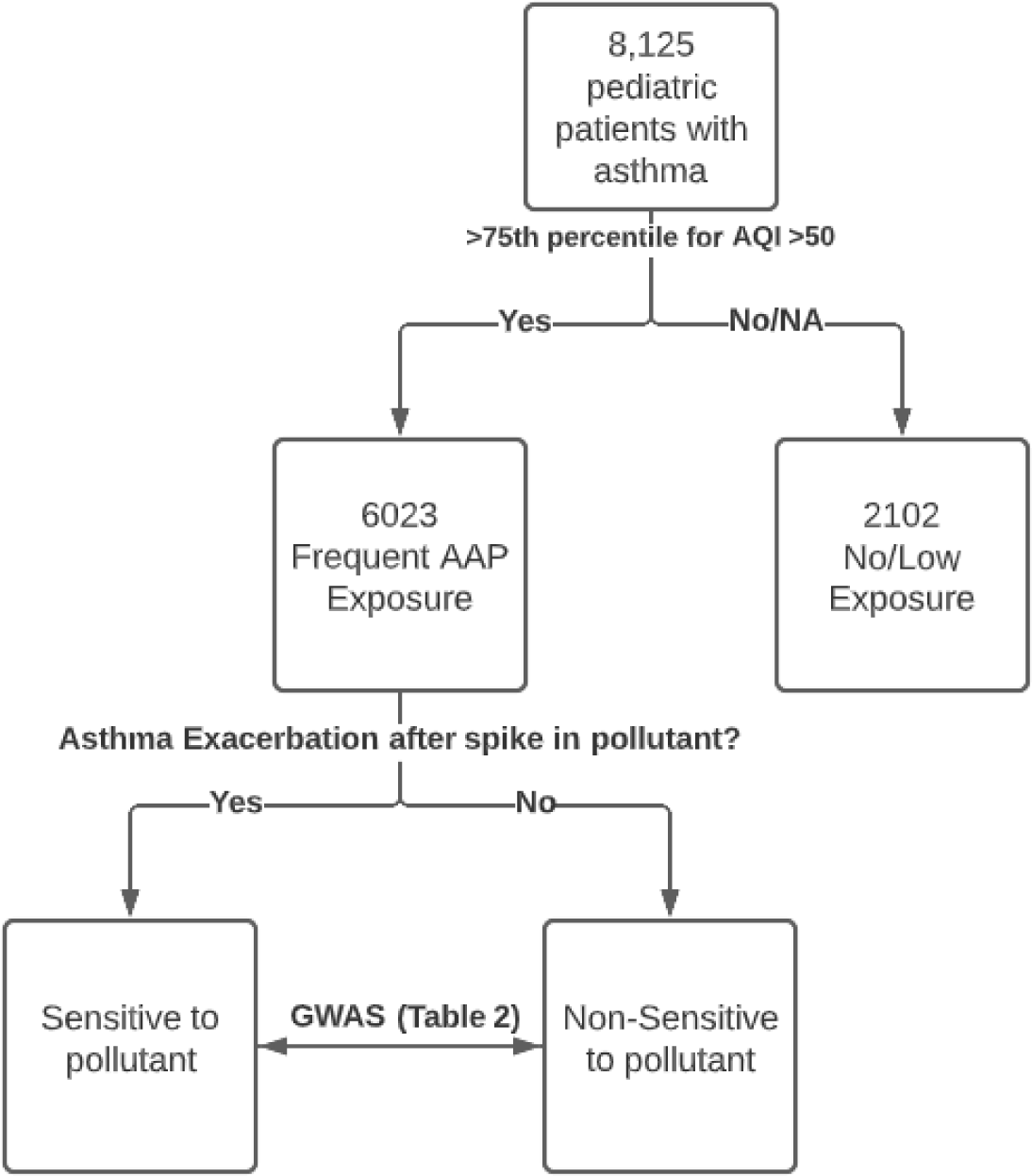
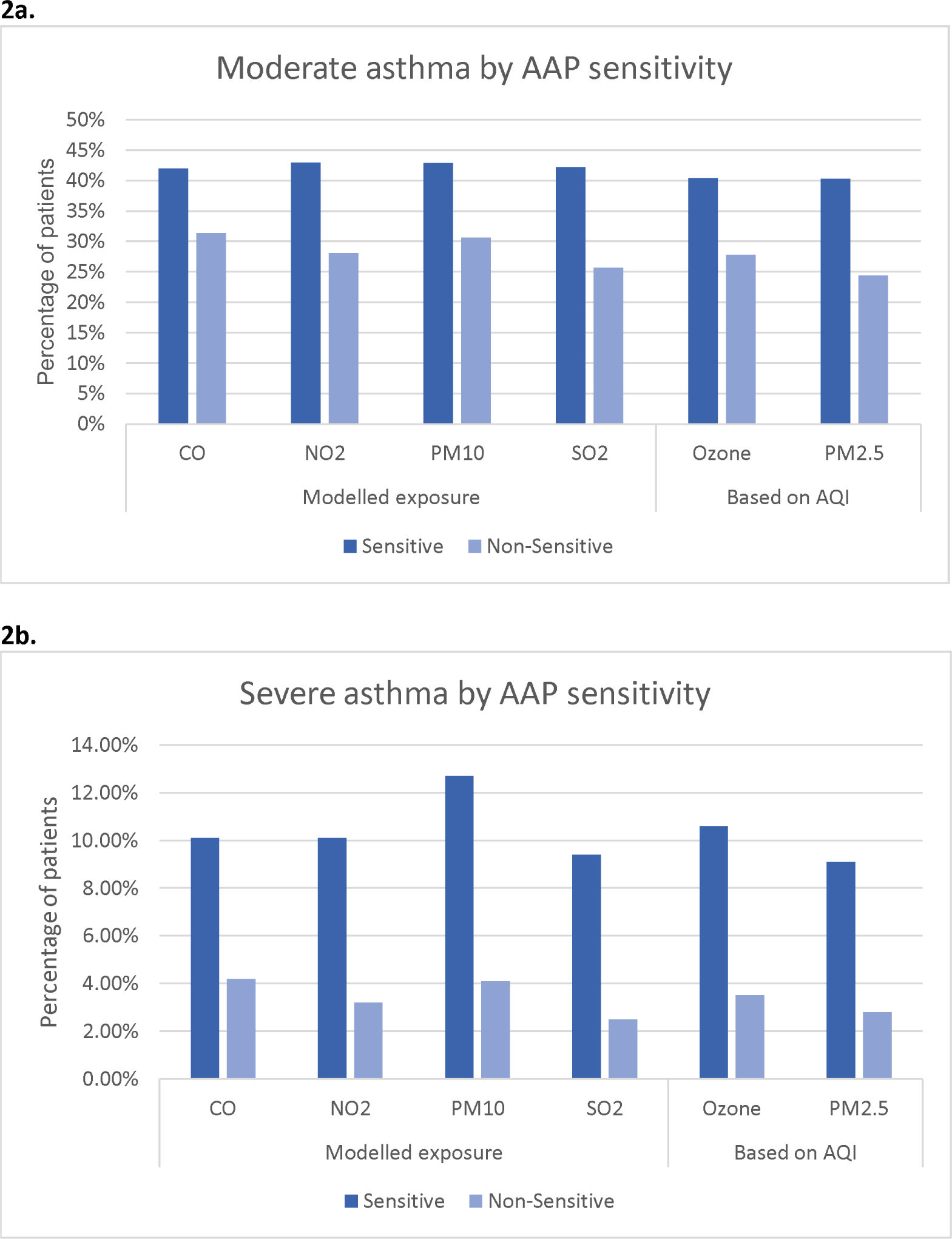
Comparison of moderate and severe asthma by AAP sensitivity status. The y-axis depicts the percentage of all patients exposed to AAP with sensitivity status available for the presented pollutant that have moderate (**2a.**) or severe (**2b.**) asthma.

AAP sensitive patients were compared to AAP non-sensitive patients by AAP component and ancestry. The African ancestry cohort was the only one large enough to complete genetic analysis. In our genetically determined African ancestry cohort: 850 CO sensitive patients were compared to 1306 CO non-sensitive patients, 1001 NO2 sensitive patients were compared to 1825 NO2 non-sensitive patients, 985 Ozone sensitive patients were compared to 2643 Ozone non-sensitive patients, 400 PM10 sensitive patients were compared to 1182 PM10 non-sensitive patients, 1462 PM2.5 sensitive patients were compared to 2408 PM2.5 non-sensitive patients, and 1319 SO2 sensitive patients were compared to 1585 SO2 non-sensitive patients. Seven single nucleotide polymorphisms (SNPs) met the genome wide significance threshold for association with AAP sensitivity (**Table 2**).

**Table 2.** Overview of SNPs significantly associated with AAP sensitivity by ancestry and pollutant. Presented odds ratios (OR) reflect odds of sensitivity to associated pollutant. The implicated gene was determined by the Open Targets Genetics Variant to Gene tool and most severe effect was determined using the Ensemble VEP tool. *MAF=Minor Allele Frequency

| Pollutant | OR | p-value | Implicated gene | Most severe effect | MAF* | Rs-ID |
| --- | --- | --- | --- | --- | --- | --- |
| CO | 6.58 | 1.63E-08 | RIMBP2 | Intron variant | 0.0097 | rs111970601 |
| NO2 | 4.37 | 2.90E-08 | ZFAT | Intergenic variant | 0.0117 | rs113389818 |
| Ozone | 3.49 | 1.21E-09 | ADGRL2 | Intergenic variant | 0.0203 | rs78061159 |
|  | 2.98 | 4.32E-08 | ADGRL2 | Regulatory Region Variant | 0.0211 | rs116515271 |
| PM10 | 0.58 | 4.62E-08 | CEACAM5 | Intron Variant | 0.2677 | rs11083637 |
| PM2.5 | 0.75 | 3.87E-09 | PFN2 | Intron Variant | 0.4747 | rs9836522 |
|  | 1.71 | 4.56E-08 | ITGB1 | Intergenic variant | 0.0663 | rs796841295 |

### In silico validation

Of SNP’s significantly associated with AAP sensitivity, rs111970601, rs11083637 and rs9836522 were classified as intron variants and rs116515271 was classified as a regulatory region variant using the VEP tool. Using the Open Targets Genetics variant to gene tool, the genes most likely functionally implicated by these variants are *RIMBP2*, *CEACAM4*, *PFN2*, and *ADGRL2* respectively. The other three variants were considered intergenic variants. Using the Open Targets Genetics interface, rs11083637 was found to be an eQTL for *CAECAM4*, *CAECAM21* and *RABAC1* and rs9836522 was found to be an eQTL for *PFN2* in blood based on eQTLGen data. (19) Locus plots were generated for non-intergenic variants using the LocusZoom tool (**e-Fig. 1**). (20) Finally, rs111970601 is located at a H3K4me1_Enh site in peripheral blood t-cells. (17)

### External validation

We were able to validate our findings regarding rs111970601 and rs9836522 in the All of Us cohort. (**e-Table 3**) For both SNPs, there were significant differences in pre-exacerbation AAP concentration (CO and PM2.5 respectively) in the direction that would be expected from our primary analysis. (**Fig. 3**) For both variants, this remained significant despite correction for self-reported race, ethnicity, sex at birth, and smoking status. (**Table 3**) Conversely, while there were significant differences in pre-exacerbation PM10 and Ozone for rs11083637 and rs116515271 respectively, but these differences were non-significant after correction for relevant confounders.

**Figure 3.**
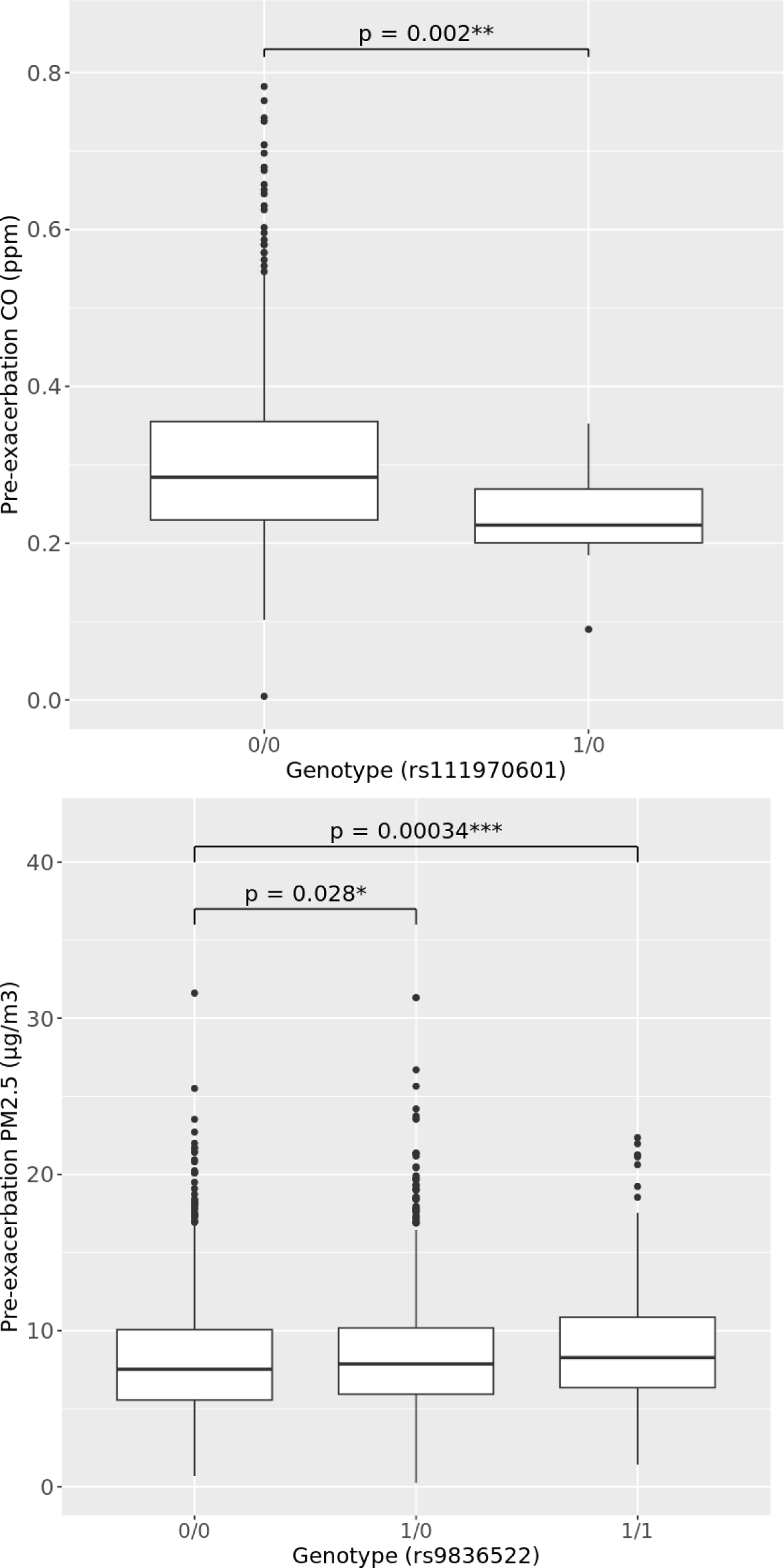
Validation - Relationship between variants and pre-exacerbation pollutant concentration in All of Us cohort. Variants that increase sensitivity are associated with lower pre-exacerbation pollutant concentration and variants that decrease sensitivity are associated with higher pre-exacerbation pollutant concentration. P-values reported here were calculated using t-tests, please refer to table 3 for p-values from regression models corrected for sex, race, ethnicity and smoking status and confidence intervals adjusted for clustering at the patient level.

**Table 3.** Validation-Relationship between variants and pre-exacerbation pollutant concentration in All of Us cohort. Presented data from linear regression models comparing mean pre-exacerbation pollutant concentration between presented genotype and 0/0 genotype. Variants that increase sensitivity are associated with lower pre-exacerbation pollutant concentration and variants that decrease sensitivity are associated with higher pre-exacerbation pollutant concentration. Models were corrected for clustering by subject, self-reported race, ethnicity, sex at birth, and lifetime history of smoking.

| Rs-ID | Pollutant | Genotype | P-value | Beta |
| --- | --- | --- | --- | --- |
| rs111970601 | CO | 1/0 | **0.0008** | -0.065 (-0.1 to - 0.03) |
| rs9836522 | PM2.5 | 1/0 | 0.28 | 0.21 (-0.13 to 0.54) |
|  |  | 1/1 | **0.02** | 0.56 (0.08 to 1.04) |

## Discussion

While previous studies have linked genetic variants and AAP to asthma incidence and reductions in FEF25-75, the presented work is the first to directly link genetic variants, AAP, and asthma exacerbations in pediatric asthma. (21, 22) The observation that variants linked with AAP sensitivity in our pediatric cohort are associated with pre-exacerbation AAP in the adult validation cohort highlight that these genetic drivers of sensitivity may have life-long consequences. The link with asthma exacerbations also offers key translational benefits. (23) Namely, since asthma exacerbations can be tracked over time, this work creates a framework to model which therapeutic interventions decrease asthma exacerbations in AAP-sensitive patients despite ongoing exposure.

In the United States, AAP exposure has been linked to race and socioeconomic status. (4, 5) We see this reconfirmed in our dataset with significantly higher rates of EMR derived ‘Black or African American’ patients living in areas with frequent occurrences of poor air quality. Given the well documented health inequities in pediatric asthma in the United States, understanding how we can mitigate the impact of environmental exposures in this population should be a priority for the research community. (24)

We have previously reported that in our cohort of pediatric patients with asthma, we can describe a subset of patients that appears sensitive to short term variations in AAP. This subgroup is more likely to have moderate or severe as opposed to mild asthma despite correction for significant confounders. (9) In the current study, we note that sensitive patients have higher rates of EMR documented allergies, family history of atopic disease, history of documented RSV and Rhinovirus infection. While some of these differences may be explained by differences in healthcare utilization, there is also evidence linking these clinical factors to AAP exposure and sensitivity. (25–28)

We identified seven SNPs that are associated with AAP sensitivity in pediatric patients with asthma. Four of these SNPs were non-intergenic with two of these validating in an external cohort. While neither of these two SNPs have been directly studied previously, the genes implicated by them have clear links to asthma and several to AAP.

First, *RIMBP2,* implicated by rs111970601, was associated with CO sensitivity. *RIMBP2* plays a role in the formation of pre-synaptic active zones shortened in mouse models of allergic asthma. (29–31) Furthermore, methylation of *RIMBP2* has been linked with lung function trajectories, and low expression of *RIMBP2* is associated with lung squamous cell carcinoma prognosis. (32, 33) Second, *PFN2* implicated by rs9836522, was associated with PM2.5 sensitivity. *PFN2* expression has been previously negatively correlated with asthma. (34) Beyond this, PFN2 is linked to small cell lung cancer metastasis and transcriptional activation of SMAD2 and SMAD3. (35, 36) This same SMAD2/3 pathway is thought to mediate the effects of PM2.5 in mice. (37) As highlighted, both *RIMBP2 and PFN2* are known to play a key role in various lung malignancies. Given the documented link between AAP exposure and lung cancer, future research should assess if sensitivity to AAP influences various diseases via common pathways. (38, 39)

Our study has some limitations. First, we only included patients that live in areas with frequent occurrences of a high AQI. While the AQI should not be confused with a detailed assessment instrument, the goal of this step was to find a group of patients at the extreme end of the exposure spectrum, ensuring that clinically relevant differences in AAP sensitivity would lead to observable differences in AAP exacerbations. In other words, if even in areas with the most frequent occurrences of poor air quality there was not an identifiable subset of patients with a relationship between timing of asthma exacerbations and spikes in air pollution, the hypothesis that differences in sensitivity lead to clinically significant differences would have been unlikely. Furthermore, we relied on indirect assessments of exposure (AQI or modelled exposure) preceding asthma exacerbation. Both approaches have their own strengths and limitations. AQI data is widely available allowing us to assign a sensitivity status to PM2.5 and Ozone to almost all patients in our cohort. By using data from days where either PM2.5 or Ozone had the highest AQI assigned to it, we tried to limit the risk that observations were driven by interactions between AAP components. As this data is collected and published directly by the EPA, it is readily available for most counties across the United States improving reproducibility of our approach. CO, NO2, SO2 and PM10 rarely have the highest AQI score, therefore, we modelled exposure to these pollutants. To this end, we relied on measuring station data and applied a squared inverse distance weighting formula to approximate exposure at the centroid of the home ZIP code. The remarkable consistency in sensitive as opposed to non-sensitive population characteristics regardless of the use of AQI or modelled data appears to support the validity of both approaches. It is worth noting prior studies using monitoring data have shown clear associations between AAP exposure and healthcare utilization in pediatric asthma despite the inexact nature of this data. (40, 41) However, recognizing the inexact nature of both approaches, we did not rely on specific cut-off values, instead, we created a relative cut-off value for every asthma exacerbation based on a reference period as outlined in the online data supplement.

Second, we used genotyping data for the GWAS part of our work and did not assess gene-gene interactions. Further exploring the genetic architecture of AAP sensitivity with whole exome and whole genome data and by modelling gene-gene interactions along the implicated pathways form key next challenges.

Our study has several unique strengths. The Center for Applied Genomics hosts the world’s largest pediatric biobank. This, together with the wide availability of EMR data for patients in our cohort and the publicly available EPA data afforded us the power needed to assess the role of genetic variants in AAP sensitivity. From a study design standpoint, our use of a temporal relationship between a short-term spike in AAP and asthma exacerbations has a key methodological advantage over assessing differences due to long term air quality. Namely, both AAP sensitive and non-sensitive patients live in the same ZIP codes limiting potential confounders based on community variables.

Our data shows that amongst pediatric patients with asthma frequently exposed to poor air quality, a subset has asthma exacerbations immediately following a spike in AAP. This subset of patients has more severe asthma. We found seven SNPs that occur more commonly in this subset of patients and were able to validate the effect of two SNPs. These SNPs implicate genes linked to asthma, AAP exposure, and lung cancer. Overall, this data suggests that there is a genetic basis for AAP sensitivity and that differences in AAP sensitivity may lead to significant clinical differences in patients exposed to AAP. To protect this subset of patients, follow-up studies will need to address how gene-gene interactions along implicated pathways and gene-environment interactions modulate the clinical consequences of AAP sensitivity and exposure.

## Data Availability

Data and code used to analyze the data available from the corresponding author upon reasonable request.

## Contributorship

Jelte Kelchtermans: Conceptualization, Investigation, Writing-Original draft preparation, Visualization.

Michael March, Frank Mentch, Yichuan Liu, Kenny Nguyen: Conceptualization, Investigation, Writing-Reviewing and Editing.

Hakon Hakonarson.: Conceptualization, Supervision, Writing-Reviewing and Editing.

## Ethics approval

This research was approved by the institutional review board of the Children’s Hospital of Philadelphia under IRB 16-013278.

## Acknowledgements

The Genotype-Tissue Expression (GTEx) Project was supported by the Common Fund of the Office of the Director of the National Institutes of Health, and by NCI, NHGRI, NHLBI, NIDA, NIMH, and NINDS. The data used for the analyses described in this manuscript were obtained from: the GTEx Portal on 03/29/23.

We gratefully acknowledge All of Us participants for their contributions, without whom this research would not have been possible. We also thank the National Institutes of Health’s All of Us Research Program for making available the participant data examined in this study.

## References

1. Pate CA, Qin X, Johnson C, Zahran HS. Asthma disparities among U.S. children and adults. The Journal of asthma : official journal of the Association for the Care of Asthma. 2023:1–10.

2. Guarnieri M, Balmes JR. Outdoor air pollution and asthma. The Lancet. 2014;383(9928):1581-92.

3. Orellano P, Quaranta N, Reynoso J, Balbi B, Vasquez J. Effect of outdoor air pollution on asthma exacerbations in children and adults: Systematic review and multilevel meta-analysis. PLOS ONE. 2017;12(3):e0174050.

4. Kravitz-Wirtz N, Crowder K, Hajat A, Sass V. THE LONG-TERM DYNAMICS OF RACIAL/ETHNIC INEQUALITY IN NEIGHBORHOOD AIR POLLUTION EXPOSURE, 1990-2009. Du Bois Review: Social Science Research on Race. 2016;13(2):237–59.

5. Clark LP, Millet DB, Marshall JD. Changes in Transportation-Related Air Pollution Exposures by Race-Ethnicity and Socioeconomic Status: Outdoor Nitrogen Dioxide in the United States in 2000 and 2010. Environmental Health Perspectives. 2017;125(9):097012.

6. Tiotiu AI, Novakova P, Nedeva D, Chong-Neto HJ, Novakova S, Steiropoulos P, et al. Impact of Air Pollution on Asthma Outcomes. International Journal of Environmental Research and Public Health. 2020;17(17):6212.

7. Stevens EL, Rosser F, Forno E, Peden D, Celedón JC. Can the effects of outdoor air pollution on asthma be mitigated? Journal of Allergy and Clinical Immunology. 2019;143(6):2016–8.e1.

8. Beasley R, Semprini A, Mitchell EA. Risk factors for asthma: is prevention possible? Lancet (London, England). 2015;386(9998):1075–85.

9. Kelchtermans J, Mentch F, Hakonarson H. Ambient air pollution sensitivity and severity of pediatric asthma. Journal of Exposure Science & Environmental Epidemiology. 2023.

10. Agency USEP. [Available from: https://aqs.epa.gov/aqsweb/airdata/download_files.html#AQI.

11. Zhou W, Nielsen JB, Fritsche LG, Dey R, Gabrielsen ME, Wolford BN, et al. Efficiently controlling for case-control imbalance and sample relatedness in large-scale genetic association studies. Nature genetics. 2018;50(9):1335–41.

12. Chang CC, Chow CC, Tellier LC, Vattikuti S, Purcell SM, Lee JJ. Second-generation PLINK: rising to the challenge of larger and richer datasets. GigaScience. 2015;4:7.

13. Ochoa D, Hercules A, Carmona M, Suveges D, Baker J, Malangone C, et al. The next-generation Open Targets Platform: reimagined, redesigned, rebuilt. Nucleic acids research. 2023;51(D1):D1353–d9.

14. McLaren W, Gil L, Hunt SE, Riat HS, Ritchie GRS, Thormann A, et al. The Ensembl Variant Effect Predictor. Genome Biology. 2016;17(1).

15. Ghoussaini M, Mountjoy E, Carmona M, Peat G, Schmidt EM, Hercules A, et al. Open Targets Genetics: systematic identification of trait-associated genes using large-scale genetics and functional genomics. Nucleic acids research. 2021;49(D1):D1311–D20.

16. Mountjoy E, Schmidt EM, Carmona M, Schwartzentruber J, Peat G, Miranda A, et al. An open approach to systematically prioritize causal variants and genes at all published human GWAS trait-associated loci. Nature genetics. 2021;53(11):1527–33.

17. Ward LD, Kellis M. HaploReg v4: systematic mining of putative causal variants, cell types, regulators and target genes for human complex traits and disease. Nucleic acids research. 2016;44(D1):D877–81.

18. Denny JC, Rutter JL, Goldstein DB, Philippakis A, Smoller JW, Jenkins G, et al. The "All of Us" Research Program. The New England journal of medicine. 2019;381(7):668–76.

19. Võsa U, Claringbould A, Westra H-J, Bonder MJ, Deelen P, Zeng B, et al. Large-scale cis- and trans-eQTL analyses identify thousands of genetic loci and polygenic scores that regulate blood gene expression. Nature genetics. 2021;53(9):1300–10.

20. Boughton AP, Welch RP, Flickinger M, VandeHaar P, Taliun D, Abecasis GR, et al. LocusZoom.js: interactive and embeddable visualization of genetic association study results. Bioinformatics (Oxford, England). 2021;37(18):3017–8.

21. Park H-W, Tantisira KG. Genetic Signatures of Asthma Exacerbation. Allergy, Asthma & Immunology Research. 2017;9(3):191.

22. Herrera-Luis E, Hernandez-Pacheco N, Vijverberg SJ, Flores C, Pino-Yanes M. Role of genomics in asthma exacerbations. Curr Opin Pulm Med. 2019;25(1):101–12.

23. Kelchtermans J, Hakonarson H. The role of gene-ambient air pollution interactions in paediatric asthma. Eur Respir Rev. 2022;31(166).

24. Forno E, Celedón JC. Health Disparities in Asthma. American Journal of Respiratory and Critical Care Medicine. 2012;185(10):1033–5.

25. Gehring U, Wijga AH, Brauer M, Fischer P, de Jongste JC, Kerkhof M, et al. Traffic-related air pollution and the development of asthma and allergies during the first 8 years of life. Am J Respir Crit Care Med. 2010;181(6):596–603.

26. Rancière F, Bougas N, Viola M, Momas I. Early Exposure to Traffic-Related Air Pollution, Respiratory Symptoms at 4 Years of Age, and Potential Effect Modification by Parental Allergy, Stressful Family Events, and Sex: A Prospective Follow-up Study of the PARIS Birth Cohort. Environmental Health Perspectives. 2017;125(4):737–45.

27. Wrotek A, Jackowska T. Molecular Mechanisms of RSV and Air Pollution Interaction: A Scoping Review. International Journal of Molecular Sciences. 2022;23(20):12704.

28. Rodrigues AF, Santos AM, Ferreira AM, Marino R, Barreira ME, Cabeda JM. Year-Long Rhinovirus Infection is Influenced by Atmospheric Conditions, Outdoor Air Virus Presence, and Immune System-Related Genetic Polymorphisms. Food and Environmental Virology. 2019;11(4):340–9.

29. Mittelstaedt T, Schoch S. Structure and evolution of RIM-BP genes: identification of a novel family member. Gene. 2007;403(1-2):70–9.

30. Wu X, Cai Q, Shen Z, Chen X, Zeng M, Du S, et al. RIM and RIM-BP Form Presynaptic Active-Zone-like Condensates via Phase Separation. Molecular cell. 2019;73(5):971–84.e5.

31. Ren M, Feng M, Long Z, Ma J, Peng X, He G. Allergic Asthma-Induced Cognitive Impairment is Alleviated by Dexamethasone. Front Pharmacol. 2021;12:680815.

32. Xu Y, She Y, Li Y, Li H, Jia Z, Jiang G, et al. Multi-omics analysis at epigenomics and transcriptomics levels reveals prognostic subtypes of lung squamous cell carcinoma. Biomed Pharmacother. 2020;125:109859.

33. Sunny SK, Zhang H, Mzayek F, Relton CL, Ring S, Henderson AJ, et al. Pre-adolescence DNA methylation is associated with lung function trajectories from pre-adolescence to adulthood. Clinical Epigenetics. 2021;13(1).

34. Roffel MP, Boudewijn IM, van Nijnatten JLL, Faiz A, Vermeulen CJ, van Oosterhout AJ, et al. Identification of asthma-associated microRNAs in bronchial biopsies. The European respiratory journal. 2022;59(3).

35. Cao Q, Liu Y, Wu Y, Hu C, Sun L, Wang J, et al. Profilin 2 promotes growth, metastasis, and angiogenesis of small cell lung cancer through cancer-derived exosomes. Aging. 2020;12(24):25981–99.

36. Tang Y-N, Ding W-Q, Guo X-J, Yuan X-W, Wang D-M, Song J-G. Epigenetic regulation of Smad2 and Smad3 by profilin-2 promotes lung cancer growth and metastasis. Nat Commun. 2015;6(1):8230.

37. Gu LZ, Sun H, Chen JH. Histone deacetylases 3 deletion restrains PM2.5-induced mice lung injury by regulating NF-κB and TGF-β/Smad2/3 signaling pathways. Biomed Pharmacother. 2017;85:756–62.

38. Wang N, Mengersen K, Tong S, Kimlin M, Zhou M, Wang L, et al. Short-term association between ambient air pollution and lung cancer mortality. Environmental Research. 2019;179:108748.

39. Xue Y, Wang L, Zhang Y, Zhao Y, Liu Y. Air pollution: A culprit of lung cancer. Journal of Hazardous Materials. 2022;434:128937.

40. Yamazaki S, Shima M, Yoda Y, Oka K, Kurosaka F, Shimizu S, et al. Exposure to air pollution and meteorological factors associated with children’s primary care visits at night due to asthma attack: case-crossover design for 3-year pooled patients. BMJ Open. 2015;5(4):e005736–e.

41. Iskandar A, Andersen ZJ, Bønnelykke K, Ellermann T, Andersen KK, Bisgaard H. Coarse and fine particles but not ultrafine particles in urban air trigger hospital admission for asthma in children. Thorax. 2012;67(3):252–7.

