## Supplementary material for "GWAS reveals Genetic Susceptibility to Air Pollution-Related Asthma Exacerbations in Children of African Ancestry": Data supplement

### Online Data Supplement

### Methods supplement

### EPA data

To assess short term variation in exposure, we used one of two approaches. For particulate matter 2.5 (PM2.5) and Ozone (O3) daily county level AQI data based on these pollutants was obtained. (E1) Patient ZIP codes were matched to counties using the reverse_zipcode function available in the zipcodeR package. (E2) ZIP codes with no AQI data available were removed. Since AQI data based on CO, NO2, PM10 and SO2 is not broadly available we modelled exposure to these pollutants using monitoring station data. Due to significant regional variation in monitoring station density and ZIP code surface area, exposure to these four pollutants was only modelled for patients living in Philadelphia County. To this end, pollution data was obtained from the Air Quality System Application Programing Interface. (E3) Data was pulled from monitoring stations in a 10- by 10-mile square centered on ZIP code centroids. The size of this square was informed by the locations of air quality monitoring stations within Philadelphia and was a compromise between the need for a small search area to preserve data validity and the requirement for a large enough area to avoid biasing results due to differential data availability. (E4) If data from multiple measuring stations was available, a squared inverse distance weighting formula was applied to model exposure. We compared AAP exposure just preceding asthma exacerbation with baseline exposure. To assess baseline exposure, we used data from two weeks prior and two weeks post the week leading up to an asthma exacerbation. This reference period was selected to be close enough to exacerbations to minimize bias based on seasonal variation while being distant enough to provide an appropriate baseline. (E5-7) Using data from this reference period, we calculated the mean and standard deviation of daily air quality index based on either O3 or PM2.5, or modelled daily pollution concentration for CO, NO2, PM10, and SO2. Patients were considered sensitive to short term spikes in a specific AAP component if they experienced at least one asthma exacerbation preceded by a spike at least two standard deviations above this mean on the day of exacerbation or three preceding days.

#### GWAS

Genotype data were generated on four major genotyping array families from Illumina (HumanHapMap550/610Q, OMNI2.5M, OmniExpress, and the GSA array). Array versions within families were merged on common SNPs and filtered for genotype missingness (geno 0.1), individual missingness (mind 0.02), and minor allele frequency (MAF >= 0.01), in that order using PLINK v1.9. (E8) Data were imputed using the TOPMed v2 reference panel on the TOPMed Imputation Server. (E9-11) Each imputed file set was filtered for imputation quality on a combination of r-squared (R2) and MAF (for SNPs with MAF >= 0.05, R2 >= 0.3 were kept; for MAF < 0.05, R2 >= 0.5 were kept). File sets were merged, and variants present in 95% of samples were retained. Ancestry was assigned based on the results of principal component analysis (PCA). PCA was performed using flashpca on approximately 2.4 million imputed SNPs with MAF > 0.05 that had been pruned for linkage disequilibrium (LD) using PLINK v1.9. (E8, E12) The first three principle components were plotted, and ancestry designation was performed by comparison to the reference genotypes from the HapMap consortium. (E13) After splitting of ancestries, ancestry-specific PCAs were performed using SNPs heavily pruned for (LD) and filtered for MAF >= 0.05. The African ancestry-specific PCA contained approximately 180,000 variants and the European ancestry-specific PCA included approximately 130,000 variants.

### Supplementary tables

|  | **Frequent AAP exposure** | **No/Low exposure** | **P value** |
| --- | --- | --- | --- |
| Number of patients | 6023 | 2102 |  |
| Sex = Male (%) | 3406 (56.5) | 1216 (57.8) | 0.312 |
| Diagnosis (%) |  |  | **<0.001** |
| Not Specified | 381 (6.3) | 496 (23.6) |  |
| Mild Asthma | 3512 (58.3) | 1056 (50.2) |  |
| Moderate Asthma | 1822 (30.3) | 448 (21.3) |  |
| Severe Asthma | 308 (5.1) | 102 (4.9) |  |
| Documented allergy = Yes (%) | 4894 (81.3) | 1734 (82.5) | 0.220 |
| Number of Exacerbations (mean (SD)) | 4.25 (5.64) | 4.17 (6.58) | 0.586 |
| Race – As documented in EMR (%) |  |  | **<0.001** |
| Asian American | 86 (1.4) | 30 (1.4) |  |
| Black or African American | 4596 (76.3) | 384 (18.3) |  |
| White | 961 (16.0) | 1476 (70.2) |  |
| Other Or missing | 380 (6.3) | 212 (10.1) |  |
| Family History of atopic disease = Yes (%) | 4351 (72.2) | 1143 (54.4) | **<0.001** |
| Obese = Yes (%) | 2030 (33.7) | 435 (20.7) | **<0.001** |
| History of RSV = Yes (%) | 603 (10.0) | 101 (4.8) | **<0.001** |
| History of Rhinovirus = Yes (%) | 620 (10.3) | 161 (7.7) | **<0.001** |
| Gestational Age (%) |  |  | 0.565 |
| Extremely preterm | 93 (1.5) | 34 (1.6) |  |
| Very preterm | 153 (2.5) | 50 (2.4) |  |
| Late preterm | 584 (9.7) | 230 (10.9) |  |
| Early term | 805 (13.4) | 272 (12.9) |  |
| Full term | 4388 (72.9) | 1516 (72.1) |  |
| Household Income - USD (mean (SD)) | 40,246 (21,376) | 75,175 (23,296) | **<0.001** |

**e-Table 1.** Comparison between included and excluded patients. P-values reported are between different pollution categories and were calculated using a chi-square test for categorical variables (with continuity correction) and ANOVA for continuous variables. P-values below 0.05 are bolded.

|  | **Modelled exposure** | | | | | |
| --- | --- | --- | --- | --- | --- | --- |
|  | **CO** | | | **NO2** | | |
|  | **Non-Sensitive** | **Sensitive** | **P-value** | **Non-Sensitive** | **Sensitive** | **P-value** |
| Number of patients | 1977 | 1282 |  | 2666 | 1471 |  |
| Sex = Male (%) | 1127 (57.0) | 728 (56.8) | 0.930 | 1486 (55.7) | 857 (58.3) | 0.125 |
| Diagnosis (%) |  |  | **<0.001** |  |  | **<0.001** |
| Not Specified | 78 (3.9) | 42 (3.3) |  | 143 (5.4) | 39 (2.7) |  |
| Mild Asthma | 1196 (60.5) | 572 (44.6) |  | 1689 (63.4) | 651 (44.3) |  |
| Moderate Asthma | 620 (31.4) | 538 (42.0) |  | 749 (28.1) | 632 (43.0) |  |
| Severe Asthma | 83 (4.2) | 130 (10.1) |  | 85 (3.2) | 149 (10.1) |  |
| Documented allergy = Yes (%) | 1594 (80.6) | 1088 (84.9) | **0.002** | 2135 (80.1) | 1247 (84.8) | **<0.001** |
| Race (%) |  |  | 0.693 |  |  | 0.800 |
| Asian American | 27 (1.4) | 23 (1.8) |  | 32 (1.2) | 22 (1.5) |  |
| Black or African American | 1640 (83.0) | 1047 (81.7) |  | 2260 (84.8) | 1253 (85.2) |  |
| White | 177 (9.0) | 122 (9.5) |  | 210 (7.9) | 111 (7.5) |  |
| Other or missing | 133 (6.7) | 90 (7.0) |  | 164 (6.2) | 85 (5.8) |  |
| Family History of atopic disease = Yes (%) | 1444 (73.0) | 980 (76.4) | **0.033** | 1904 (71.4) | 1173 (79.7) | **<0.001** |
| Obese = Yes (%) | 683 (34.5) | 484 (37.8) | 0.068 | 945 (35.4) | 541 (36.8) | 0.412 |
| Gestational Age (%) |  |  | **<0.001** |  |  | 0.198 |
| Extremely preterm | 25 (1.3) | 27 (2.1) |  | 36 (1.4) | 23 (1.6) |  |
| Very preterm | 47 (2.4) | 39 (3.0) |  | 60 (2.3) | 48 (3.3) |  |
| Late preterm | 181 (9.2) | 147 (11.5) |  | 261 (9.8) | 152 (10.3) |  |
| Early term | 239 (12.1) | 198 (15.4) |  | 352 (13.2) | 209 (14.2) |  |
| Full term | 1485 (75.1) | 871 (67.9) |  | 1957 (73.4) | 1039 (70.6) |  |
| Mean household income (mean (SD)) | 32,291 (10,587) | 31,990 (10,923) | 0.435 | 31,739 (9,998) | 31,679 (9,688) | 0.850 |

**e-Table 2.** Comparison between sensitive and non-sensitive patients by AAP component. P-values were calculated using a chi-square test for categorical variables (with continuity correction) and ANOVA for continuous variables. P-values below 0.05 are bolded.

|  | **Modelled exposure** | | | | | |
| --- | --- | --- | --- | --- | --- | --- |
|  | **PM10** | | | **SO2** | | |
|  | **Non-Sensitive** | **Sensitive** | **P-value** | **Non-Sensitive** | **Sensitive** | **P-value** |
| Number of patients | 1823 | 671 |  | 2323 | 1905 |  |
| Sex = Male (%) | 1014 (55.6) | 406 (60.5) | **0.032** | 1313 (56.5) | 1078 (56.6) | 0.990 |
| Diagnosis (%) |  |  | **<0.001** |  |  | **<0.001** |
| Not Specified | 83 (4.6) | 25 (3.7) |  | 140 (6.0) | 52 (2.7) |  |
| Mild Asthma | 1109 (60.8) | 273 (40.7) |  | 1527 (65.7) | 871 (45.7) |  |
| Moderate Asthma | 557 (30.6) | 288 (42.9) |  | 597 (25.7) | 803 (42.2) |  |
| Severe Asthma | 74 (4.1) | 85 (12.7) |  | 59 (2.5) | 179 (9.4) |  |
| Documented allergy = Yes (%) | 1487 (81.6) | 575 (85.7) | **0.019** | 1822 (78.4) | 1648 (86.5) | **<0.001** |
| Race (%) |  |  | **0.004** |  |  | 0.816 |
| Asian American | 30 (1.6) | 15 (2.2) |  | 28 (1.2) | 25 (1.3) |  |
| Black or African American | 1477 (81.0) | 504 (75.1) |  | 1988 (85.6) | 1624 (85.2) |  |
| White | 191 (10.5) | 80 (11.9) |  | 166 (7.1) | 148 (7.8) |  |
| Other or missing | 125 (6.9) | 72 (10.7) |  | 141 (6.1) | 108 (5.7) |  |
| Family History of atopic disease = Yes (%) | 1271 (69.7) | 519 (77.3) | **<0.001** | 1640 (70.6) | 1498 (78.6) | **<0.001** |
| Obese = Yes (%) | 652 (35.8) | 240 (35.8) | 1.000 | 776 (33.4) | 741 (38.9) | **<0.001** |
| Gestational Age (%) |  |  | **0.009** |  |  | **<0.001** |
| Extremely preterm | 25 (1.4) | 14 (2.1) |  | 29 (1.2) | 32 (1.7) |  |
| Very preterm | 45 (2.5) | 21 (3.1) |  | 52 (2.2) | 56 (2.9) |  |
| Late preterm | 163 (8.9) | 83 (12.4) |  | 203 (8.7) | 213 (11.2) |  |
| Early term | 213 (11.7) | 93 (13.9) |  | 278 (12.0) | 292 (15.3) |  |
| Full term | 1377 (75.5) | 460 (68.6) |  | 1761 (75.8) | 1312 (68.9) |  |
| Mean household income (mean (SD)) | 32,341 (11,312) | 31,898 (11,257) | 0.386 | 32,128 (9,807) | 31,646 (9,872) | 0.112 |

**e-Table 2. (Cont.).** Comparison between sensitive and non-sensitive patients by AAP component. P-values reported were calculated using a chi-square test for categorical variables (with continuity correction) and ANOVA for continuous variables. P-values below 0.05 are bolded.

|  | **Based on AQI** | | | | | |
| --- | --- | --- | --- | --- | --- | --- |
|  | **Ozone** | | | **PM2.5** | | |
|  | **Non-Sensitive** | **Sensitive** | **P-value** | **Non-Sensitive** | **Sensitive** | **P-value** |
| Number of patients | 4113 | 1504 |  | 3800 | 2219 |  |
| Sex = Male (%) | 2340 (56.9) | 854 (56.8) | 0.965 | 2121 (55.8) | 1283 (57.8) | 0.137 |
| Diagnosis (%) |  |  | **<0.001** |  |  | **<0.001** |
| Not Specified | 281 (6.8) | 67 (4.5) |  | 278 (7.3) | 103 (4.6) |  |
| Mild Asthma | 2544 (61.9) | 671 (44.6) |  | 2491 (65.6) | 1018 (45.9) |  |
| Moderate Asthma | 1143 (27.8) | 607 (40.4) |  | 926 (24.4) | 895 (40.3) |  |
| Severe Asthma | 145 (3.5) | 159 (10.6) |  | 105 (2.8) | 203 (9.1) |  |
| Documented allergy = Yes (%) | 3292 (80.0) | 1304 (86.7) | **<0.001** | 3014 (79.3) | 1877 (84.6) | **<0.001** |
| Race (%) |  |  | 0.735 |  |  | **0.019** |
| Asian American | 63 (1.5) | 20 (1.3) |  | 48 (1.3) | 38 (1.7) |  |
| Black or African American | 3148 (76.5) | 1147 (76.3) |  | 2862 (75.3) | 1733 (78.1) |  |
| White | 640 (15.6) | 248 (16.5) |  | 637 (16.8) | 321 (14.5) |  |
| Other or missing | 262 (6.4) | 89 (5.9) |  | 253 (6.7) | 127 (5.7) |  |
| Family History of atopic disease = Yes (%) | 2921 (71.0) | 1163 (77.3) | **<0.001** | 2625 (69.1) | 1726 (77.8) | **<0.001** |
| Obese = Yes (%) | 1386 (33.7) | 522 (34.7) | 0.499 | 1210 (31.8) | 819 (36.9) | **<0.001** |
| Gestational Age (%) |  |  | 0.085 |  |  | **0.043** |
| Extremely preterm | 59 (1.4) | 33 (2.2) |  | 58 (1.5) | 35 (1.6) |  |
| Very preterm | 103 (2.5) | 42 (2.8) |  | 90 (2.4) | 63 (2.8) |  |
| Late preterm | 389 (9.5) | 153 (10.2) |  | 352 (9.3) | 232 (10.5) |  |
| Early term | 541 (13.2) | 220 (14.6) |  | 481 (12.7) | 324 (14.6) |  |
| Full term | 3021 (73.5) | 1056 (70.2) |  | 2819 (74.2) | 1565 (70.5) |  |
| Mean household income (mean (SD)) | 39,534 (20,589) | 41,245 (22,579) | **0.007** | 41,224 (22,437) | 38,531 (19,287) | **<0.001** |

**e-Table 2. (Cont.).** Comparison between sensitive and non-sensitive patients by AAP component. P-values reported were calculated using a chi-square test for categorical variables (with continuity correction) and ANOVA for continuous variables. P-values below 0.05 are bolded.

|  | **rs111970601** | **rs11083637** | **rs9836522** | **rs116515271** |
| --- | --- | --- | --- | --- |
| Number of patients | 315 | 1108 | 1505 | 1483 |
| Number of asthma exacerbations | 787 | 2365 | 3378 | 3331 |
| Genotype (%) |  |  |  |  |
| 1/0 | <20 | 476 (43) | 649 (43) | <20 |
| 1/1 | 0 | 90 (8) | 201 (13) | 0 |
| Self-reported Race (%) |  |  |  |  |
| Black or African American | 258 (82) | 233 (21) | 287 (19) | 281 (19) |
| White | 0 | 673 (61) | 888 (59) | 880 (59) |
| Other or missing | 57 (18) | 202 (18) | 330 (22) | 322 (22) |
| History of smoking – Yes (%) | 127 (40) | 462 (42) | 598 (40) | 596 (40) |
| Self-reported Ethnicity (%) |  |  |  |  |
| Hispanic or Latino | <35 (11) | <150 (13) | <260 (17) | <250 (17) |
| Not Hispanic or Latino | 261 (83) | 918 (83) | 1192 (79) | 1175 (79) |
| Sex at birth (%) |  |  |  |  |
| Female | 264 (84) | 863 (78) | 1174 (78) | 1157 (78) |
| Male | <50 (15) | <240 (21) | <320 (21) | <310 (21) |
| Age (years) – Mean (Sd) | 51 (13) | 56 (14) | 56 (14) | 56 (14) |

**e-Table 3.** Overview of validation cohort per SNP. As per All of Us policies, categories were collapsed and exact numbers hidden if sub-groups contained fewer than 20 subjects.

### e-Figure Legends

**e-Figure 1.** **Regional LocusZoom plots of regions flanking those non-intergenic SNPs significantly associated with AAP sensitivity.** Plots were generated using the LocalZoom tool, Plink 1.9 was used to calculate linkage disequilibrium information relative to reference variants (purple diamonds).

### Supplementary references

E1.	Agency USEP. [Available from: <https://aqs.epa.gov/aqsweb/airdata/download_files.html#AQI>.

E2.	Rozzi GC. zipcodeR: Advancing the analysis of spatial data at the ZIP code level in R. Software Impacts. 2021;9:100099.

E3.	Kelchtermans J, Mentch F, Hakonarson H, editors. Facilitating Case-Crossover Studies Using Environmental Protection Agency Data2022: American Thoracic Society.

E4.	2021-2022 Air Monitoring Network Plan. In: City of Philadelphia DoPH, Air Management Services, editor. 2021.

E5.	Yamazaki S, Shima M, Yoda Y, Oka K, Kurosaka F, Shimizu S, et al. Exposure to air pollution and meteorological factors associated with children's primary care visits at night due to asthma attack: case-crossover design for 3-year pooled patients. BMJ Open. 2015;5(4):e005736-e.

E6.	Yu-Ni H, Fu-Jen C, Ming-Ta T, Chih-Min T, Po-Chun C, Chi-Yung C. Fine particulate matter constituents associated with emergency room visits for pediatric asthma: a time-stratified case–crossover study in an urban area. 2021.

E7.	Peng RD, Dominici F, Pastor-Barriuso R, Zeger SL, Samet JM. Seasonal analyses of air pollution and mortality in 100 US cities. Am J Epidemiol. 2005;161(6):585-94.

E8.	Chang CC, Chow CC, Tellier LC, Vattikuti S, Purcell SM, Lee JJ. Second-generation PLINK: rising to the challenge of larger and richer datasets. GigaScience. 2015;4:7.

E9.	Das S, Forer L, Schönherr S, Sidore C, Locke AE, Kwong A, et al. Next-generation genotype imputation service and methods. Nature genetics. 2016;48(10):1284-7.

E10.	Fuchsberger C, Abecasis GR, Hinds DA. minimac2: faster genotype imputation. Bioinformatics (Oxford, England). 2015;31(5):782-4.

E11.	Taliun D, Harris DN, Kessler MD, Carlson J, Szpiech ZA, Torres R, et al. Sequencing of 53,831 diverse genomes from the NHLBI TOPMed Program. Nature. 2021;590(7845):290-9.

E12.	Abraham G, Inouye M. Fast principal component analysis of large-scale genome-wide data. PLoS One. 2014;9(4):e93766.

E13.	Altshuler DM, Gibbs RA, Peltonen L, Altshuler DM, Gibbs RA, Peltonen L, et al. Integrating common and rare genetic variation in diverse human populations. Nature. 2010;467(7311):52-8.
